# Laboratory validation of a clinical metagenomic next-generation sequencing assay for respiratory virus detection and discovery

**DOI:** 10.1101/2024.02.28.24303521

**Authors:** Jessica Karielle Tan, Venice Servellita, Doug Stryke, Emily Kelly, Jessica Streithorst, Nanami Sumimoto, Abiodun Foresythe, Hee Jae Huh, Jenny Nguyen, Miriam Oseguera, Noah Brazer, Jack Tang, Danielle Ingebrigtsen, Becky Fung, Helen Reyes, Melissa Hillberg, Alice Chen, Hugo Guevara, Shigeo Yagi, Christina Morales, Debra A. Wadford, Peter M. Mourani, Charles R. Langelier, Mikael de Lorenzi-Tognon, Patrick Benoit, Charles Y. Chiu

## Abstract

Tools for rapid identification of novel and/or emerging viruses are urgently needed for clinical diagnosis of unexplained infections and pandemic preparedness. Here we developed and clinically validated a largely automated metagenomic next-generation sequencing (mNGS) assay for agnostic detection of respiratory viral pathogens from nasopharyngeal swab, bronchoalveolar lavage and tracheal aspirate samples in <24 hours. The mNGS assay achieved mean limits of detection of 543 copies/mL, viral load quantification with 100% linearity, and 93.6% sensitivity, 93.8% specificity, and 93.7% accuracy compared to gold-standard clinical multiplex RT-PCR. Performance increased to 97.9% overall predictive agreement after discrepancy testing and clinical adjudication, which was superior to that of RT-PCR (95.0% agreement). To enable discovery of novel, sequence-divergent human viruses with pandemic potential, *de novo* assembly and translated nucleotide algorithms were incorporated into the automated SURPI+ computational pipeline used by the mNGS assay for pathogen detection. Using *in silico* analysis, we showed after removal of all human viral sequences from the reference database that 70 (100%) of 70 representative human viral pathogens could still be identified based on homology to related animal or plant viruses. Our assay, which was granted breakthrough device designation from the US Food and Drug Administration (FDA) in August of 2023, demonstrates the feasibility of routine mNGS testing in clinical and public health laboratories, thus enabling a robust and rapid response to the next viral respiratory pandemic.

## Introduction

Respiratory infections are among the most common infections globally and are associated with significant morbidity and mortality^1–3^. Despite their importance, half of adult patients hospitalized in the United States with community-acquired pneumonia, which is most commonly caused by respiratory viruses, have no causative pathogen identified^2–5^. Respiratory infections caused by viruses can be especially challenging to diagnose because of the diversity of potential agents^6–8^. In particular, emerging pandemic viruses represent an unpredictable threat which traditional diagnostic tools such as nucleic acid amplification tests have not been designed to detect^9^. The importance of unbiased assays for rapid identification of viral pathogens, especially those with sequence-divergent genomes, became evident during the discovery of SARS-CoV-2^10,11^

Metagenomic next-generation sequencing (mNGS) has emerged as an attractive diagnostic method for identifying causative agents in unexplained infections as it provides a comprehensive and agnostic approach by which all potential pathogens can be identified in a single assay without the need for specific primers and probes^12,13^. mNGS has been used for broadly diagnosing infections, whether viral, bacterial, fungal, or parasitic, from multiple specimen types^14–16^, and its clinical utility has been demonstrated for neurological and bloodstream infections^16–18^.

However, despite the favorable performance of mNGS testing demonstrated by multiple studies, general adoption of mNGS technologies in clinical microbiology laboratories has been hindered by high costs, complex protocols, lack of automation, insufficient standardization of bioinformatic pipelines, prolonged turnaround times (24-72 hours), lack for regulatory guidelines for clinical validation, and overall lower sensitivity for detection of common pathogens relative to targeted approaches such as polymerase chain reaction (PCR) assays^19^.

Here we describe the development, optimization, and clinical validation of a streamlined and largely automated mNGS laboratory-developed test (LDT) with a sample-to-result turnaround time of less than 24 hours for identification of common as well as unexpected and/or novel viral respiratory pathogens. The computational SURPI+ pipeline used by the mNGS assay was modified to provide enhanced analysis capabilities, including viral load quantification, incorporation of curated reference genome databases such as FDA dAtabase for Reference Grade micrObial Sequences (FDA-ARGOS), and sensitive identification of novel, sequence-divergent viruses by *de novo* assembly and translated nucleotide alignment. We comprehensively evaluated assay performance metrics, including limits of detection, linearity, precision, inclusivity and exclusivity, contamination, interference, matrix effect, stability, accuracy, and capacity to detect novel viruses.

## Results

### Development and Optimization of an mNGS Assay for Detection of Viral Respiratory Pathogens

We developed an mNGS assay for the detection of viral pathogens from respiratory secretions, including nasopharyngeal swab (NPS) samples, bronchoalveolar lavage (BAL) fluid and tracheal aspirate **(Figure 1)**. The sample preparation protocol was optimized to maximize sensitivity and decrease assay turnaround time. We tested different combinations of centrifugation, heat, and addition of a DNA/RNA stabilization medium prior to total nucleic acid extraction and found that centrifugation alone produced the highest yield of detected viral reads. To decrease turnaround times, we used a 15-minute protocol for human rRNA depletion and reduced incubation times for the reverse transcription and second-strand cDNA synthesis steps to 15 and 9 minutes, respectively. The final assay used 450 μL of sample input volume and consisted of the following steps: (1) centrifugation (∼15 min), total nucleic acid extraction and DNase treatment for isolation of total RNA (∼1 hr), (2) cDNA synthesis with ribosomal RNA (rRNA) depletion (∼1 hr), (3) barcoded adapter ligation, library PCR amplification and purification on an automated instrument (∼6.5 hr), (4) library pooling (∼5 min), (5) Illumina (San Diego, CA) sequencing (5 or 13 hr, depending on whether a MiniSeq or NextSeq sequencer is used), and (6) bioinformatics analysis for viral detection and quantification using the SURPI+ pipeline (∼0.5 hr). Overall sample-to-answer assay turnaround time was 14 − 22 hours. We used MS2 phage and External RNA Controls Consortium (ERCC) RNA Spike-In Mix (Invitrogen, Waltham, MA) added into each sample as internal qualitative and quantitative controls, respectively. The MS2 phage and ERCC sequencing results were also used to evaluate and interpret the background level in the sample, generally originating from the human host **(Supplementary Tables 1and 2)**. A commercial reference panel (Accuplex Panel, SeraCare, Milford, MA) consisting of quantified SARS-CoV-2, influenza A, influenza B, and respiratory syncytial virus (RSV) was spiked into pooled virus-negative nasopharyngeal swab matrix (see Methods for details) as an external positive control (PC) for the assay, with the negative matrix serving as an external negative control (NC).

**Figure 1.**
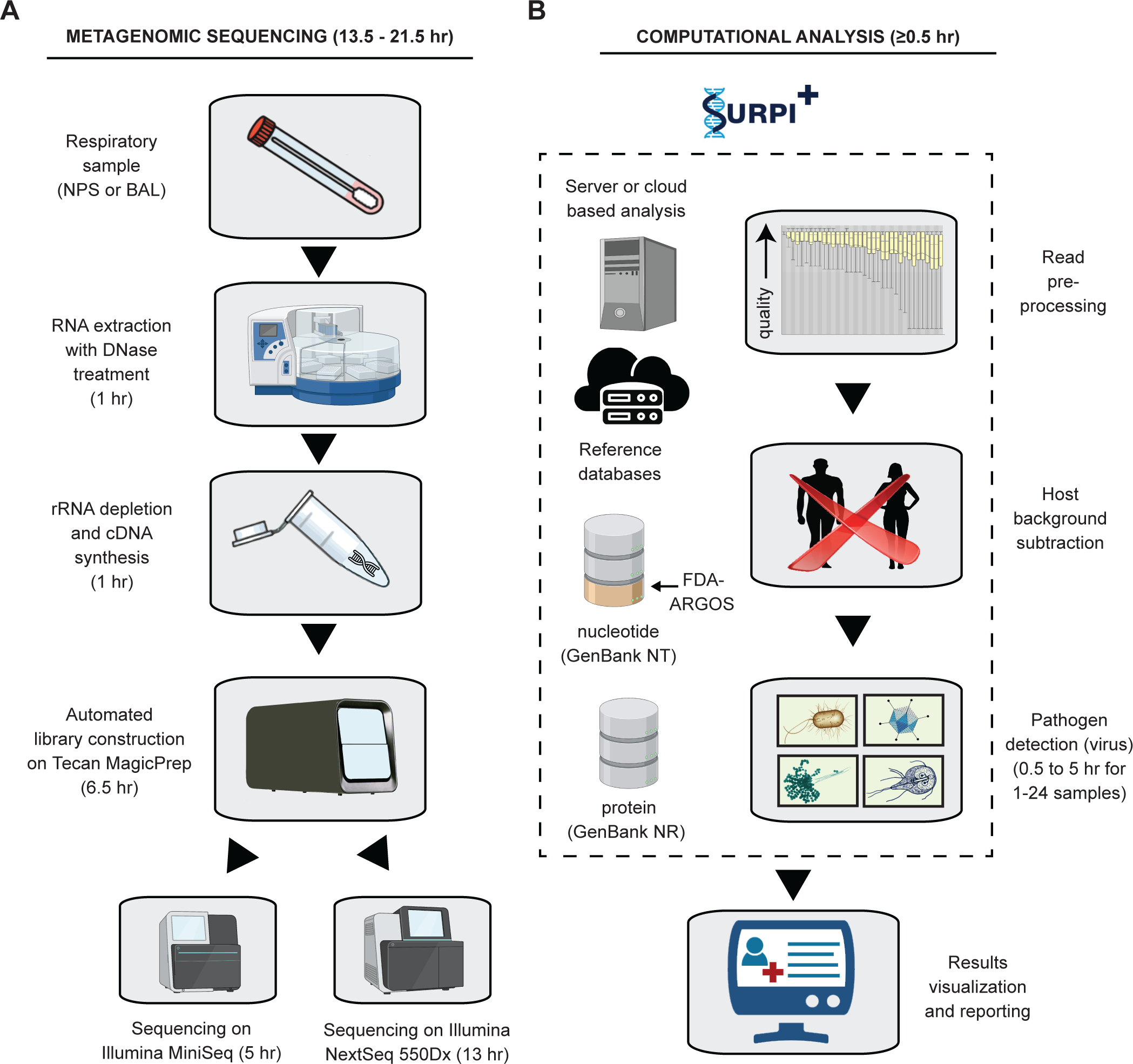
Schematic of the mNGS assay workflow. **(A)** RNA from respiratory samples is extracted and treated with DNase. Internal control is added to assess human background during sequencing. Human rRNA is depleted during cDNA synthesis. Libraries are generated on the automated Tecan MagicPrep NGS instrument. Libraries are normalized, pooled, and loaded onto the sequencer. **(B)** Sequences are processed using SURPI+ software for alignment and classification. Reads are preprocessed by trimming of adapters and removal of low-quality/low-complexity sequences, followed by computational subtraction of human reads. SURPI+ pipeline has been modified (Figure 2) to analyze only viral metagenomic data. Reads are mapped to the closest matched genome to identify non overlapping regions using NCBI GenBank and FDA-ARGOS database. To aid in analysis, automated result summaries, heat maps of raw/normalized read counts, and coverage/pairwise identity plots are generated for clinical interpretation. Total turnaround time is between 14 and 22 hours depending on type of sequencer used.

The SURPI+ computational pipeline, run as a container on either a server or cloud, was used for the identification of viral respiratory pathogens from mNGS data^20,21^. Three enhancements were made (**Figure 2A**). First, we added the capability for viral load quantification using the PC and a standard curve generated for each sample from the ERCC reads. Second, a custom algorithm consisting of *de novo* assembly of metagenomic reads and translated nucleotide, or amino acid, alignment of the reads to a viral protein database was developed to enable detection of novel, sequence-divergent viruses. Third, “tagging” of taxonomic entries in the SURPI+ database was incorporated to allow inclusion of curated viral reference genomes, such as those deposited in the FDA-ARGOS database^22^, for virus identification by alignment and results reporting.

**Figure 2.**
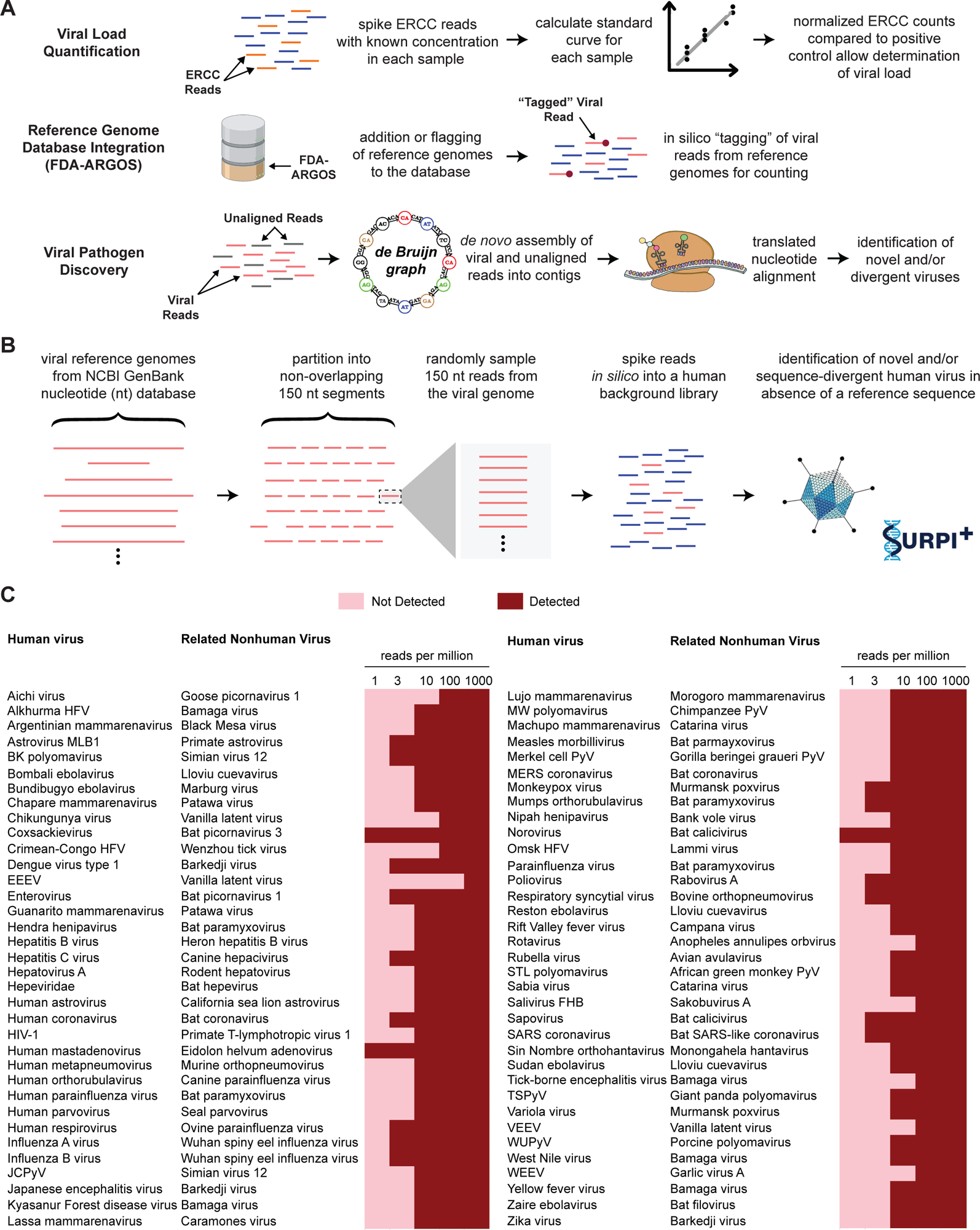
Enhancements to the SURPI+ Bioinformatics Pipeline for Pathogen Identification. **(A)** Schematic diagram of SURPI+ software modifications to enable reporting of the estimated viral load using a quantitative internal spiked control (ERCC). A standard curve is generated for each sample using the normalized ERCC results and absolute quantification by comparison of the ERCC data with the external PC. Translated viral reads and nucleotides are aligned to reference sequences in GenBankNRand GenBank NT, respectively. Each read is annotated with the lowest taxonomic rank that comprises a given threshold fraction of the read’s total alignments to that rank, including alignments to the reference-grade sequences in the FDA-ARGOS database. Number of reads mapped to GenBank NT and FDA-ARGOS is shown on the clinical report. De novo viral genome assembly and translated nucleotide (amino acid) alignments are done using the SPAdes and DIAMOND algorithms, respectively. After assembly of contigs, both the contigs and the raw unaligned reads are then processed through DIAMOND to identify sequences that may correspond to novel, highly divergent viruses that may be the basis for a newly emerging pandemic. **(B)** Representative viral reference genomes corresponding to outbreak viruses of clinical and public health significance with pandemic potential are retrieved from the NCBI GenBank database, partitioned into non-overlapping segments, and then randomly sampled and spiked *in silico* into a negative nasal swab matrix sequencing library. A higher-level set of taxonomic identifiers (species, genus, and/or family) corresponding to these viruses are removed from the SURPI+ reference dataset and the simulated sequencing file is analyzed using both the original and “restricted reference” databases. **(C)** Viruses can be detected using the modified SURPI+ pipeline despite lacking a taxonomic reference at levels down to 10-100 reads per million (RPM). Abbreviations: EEEV, Eastern equine encephalitis virus; ERCC; External RNA Controls Consortium; FDA-ARGOS, FDA dAtabase for Reference Grade micrObial Sequences; HFV, hemorrhagic fever virus; HIV, human immunodeficiency virus; JCPyV, JC polyomavirus; PC, positive control; PyV, polyomavirus; TSPyV, trichodysplasia spinulosa polyomavirus; SURPI, sequence-based ultrarapid pathogen identification; VEEV, Venezuelan equine encephalitis virus; WEEV, Western equine encephalitis virus.

Quality control metrics were based on those previously established for a validated cerebrospinal fluid mNGS assay^20^ and include a minimum of 5 million preprocessed reads per sample, >75% of data with quality score >30 (Q>30), and successful detection of the internal spiked MS2 phage control and all four respiratory viruses in the PC. A threshold criterion of ≥3 non-overlapping viral reads aligning to the target viral genome was considered a positive detection.

### Analytical Sensitivity

We adopted Clinical and Laboratory Standards Institute (CLSI) guidelines for NGS-based infectious diseases testing (MM24)^23^ and validation of multiplex nucleic acid assays (MM17)^24^ to conduct a comprehensive evaluation of assay performance metrics **(Table 1)**. To determine limits of detection (LoD), negative NPS matrix was spiked with the Accuplex Verification Panel and diluted at concentrations ranging from 5,000 to 100 copies/mL, with 10 to 40 replicates at each concentration. By 95% probit analysis, the LoD was determined for each of the four representative organisms in the panel (SARS-CoV-2, Influenza A, Influenza B, and RSV). We found LoDs ranging from 439 to 706 copies/mL for the four respiratory viruses in the positive control **(Figure 3)**. The achieved average LoD of 550 copies/mL was comparable within one log to reported LoDs from specific reverse transcription-polymerase chain reaction (RT-PCR) assays for detection of viral respiratory pathogens^25^.

**Figure 3.**
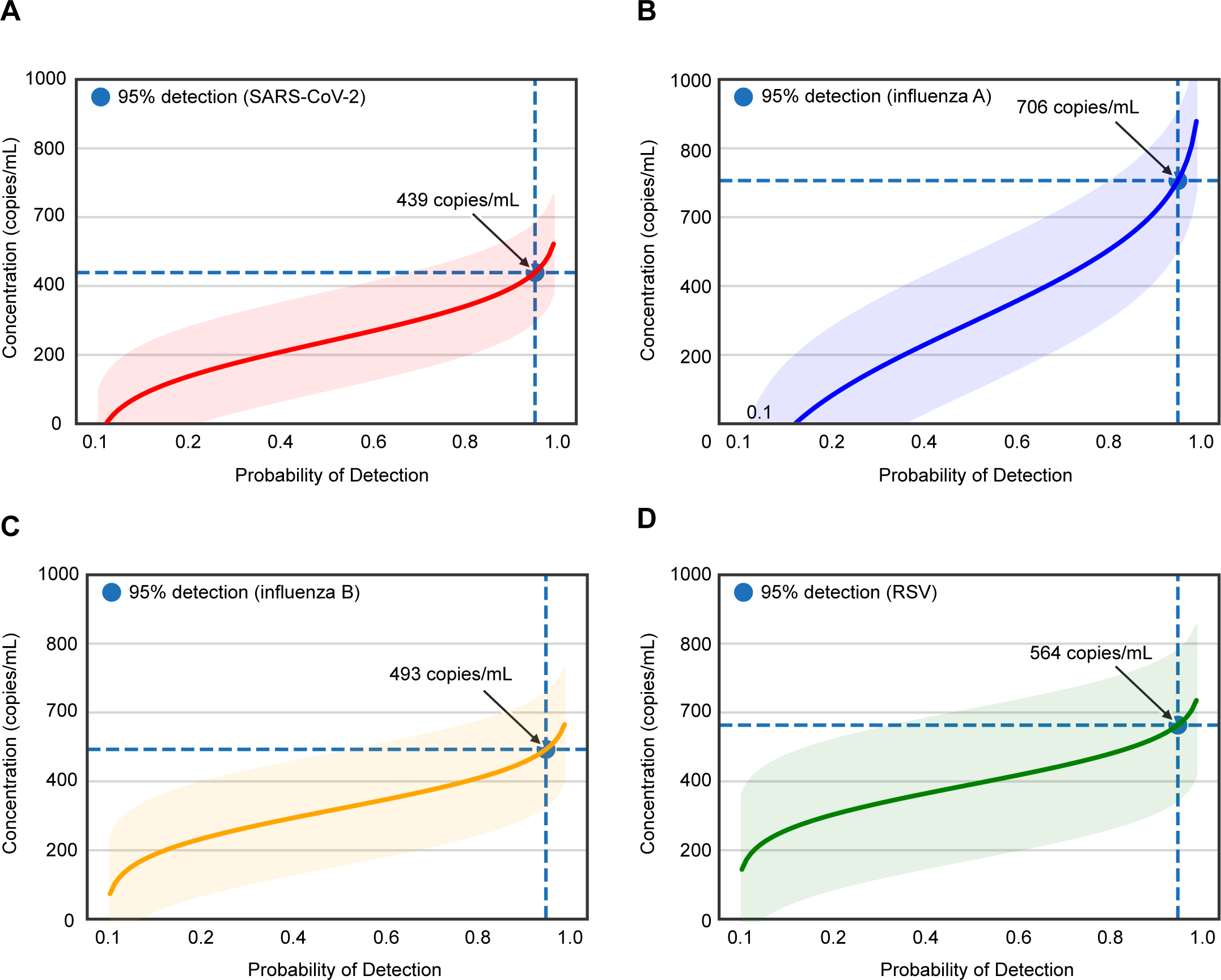
Limits of detection (LoD) study. Probit regression analysis curves plotting the viral titer in copies/mL (y-axis) against the calculated detection probability (x-axis) of (A) SARS-CoV-2, (B) Influenza A, (C) Influenza B and (D) Respiratory Syncytial Virus (RSV). The detection probability corresponding to 95% is denoted with a blue circle for each virus. Shaded areas represent the 95% confidence intervals for each curve. Probit analyses were done using Python software (version 3.7.12). Results show a LoD ranging from 439 to 706 copies/mL for the 4 respiratory viruses in the positive control.

**Table 1.** Performance characteristics of the Viral Respiratory Pathogens (VRP) mNGS assay.

| Metrics | Method | Expected target |  | Results |  |
| --- | --- | --- | --- | --- | --- |
| Limit of detection (LoD) | Detection of PC dilution by Probit analysis | <1000 copies/mL |  | Target<br>SARS-CoV-2<br>Influenza A<br>Influenza B<br>RSV | LoD<br>439 copies/mL<br>706 copies/mL<br>493 copies/mL<br>563 copies/mL |
| Linearity | Correlation of PC with assay quantification | $R^2 > 90\%$ | | $R^2 = 100\%$ | |
| Precision | Intra-Assay: PC and NC within the same run across 20 runs. | Concordance 100% EA | Log-transformed CV <10% | Concordance 100% EA | Log-transformed CV <10% |
|  | Inter-Assay: PC and NC across 20 separate runs | 100% EA | <30% | 100% EA | <30% |
| Inclusivity | Detection of viruses from diluted culture supernatant | 100% detection |  | 100% detection (17/17) |  |
|  | Detection of viruses in positive BAL/CSF diluted samples | 100% detection |  | 100% detection (11/11) |  |
| Exclusivity | Detection of viruses in a known organism mixture<sup>a</sup> | No false-positive |  | No false-positive |  |
| Contamination | Detection of cross-contamination on the sample wells | No carryover contamination |  | Cross-contamination of 0.1% between adjacent wells but no carryover contamination |  |
| Interference | Detection of PC spiked with hemolytic blood | Detection at all concentrations |  | Detection at all concentrations |  |
|  | Detection of PC spiked with Human RNA | Detection at all concentrations |  | Detection at all concentrations |  |
| | Detection of PC spiked with bacterial DNA/RNA | Detection at concentration $\leq 10^7$ cells/mL | | Detection at concentration $\leq 10^7$ cells/mL | |
|  | Detection of virus-positive overtly mucoid BAL samples | Detection in all BAL samples |  | Target detected in 13/14 (92.9%) valid sample runs |  |
| Stability | Detection of targets in samples held at 4°C for 7 days or after 3 freeze-thaw cycles | 100% concordance |  | 100% concordance |  |
| Accuracy | Detection in virus positive and negative samples (n=191) | Sensitivity > 90%<br>Specificity > 90%<br>Accuracy > 90%<br>PPA > 90%<br>NPA > 90% |  | Original testing<br>Sensitivity: 93.6%<br>Specificity: 93.8 %<br>Accuracy: 93.7 % | After discrepancy testing and clinical adjudication<br>PPA: 98.7%<br>NPA: 98.1%<br>Overall: 97.9% |
| Detection of divergent viruses | Detection by an *in silico* analysis of divergent viruses (n=70) | Sensitivity >95%<br>Specificity >95% |  | Sensitivity: 98.6%<br>Specificity: 100% |  |

(PC) Positive control consisting of 4 respiratory viruses spiked into pooled nasopharyngeal swab matrix; (IC) spiked internal control consisting of a RNA MS2 phage; (NC) Negative control; (EA) Essential agreement, (CV) Coefficient of variation, (PPA) positive percent agreement; (NPA) negative percent agreement.
<sup>a</sup>Mixture includes detectable concentrations of CMV, HIV, *Klebsiella pneumoniae*, *Streptococcus agalactiae*, *Aspergillus niger*, *Cryptococcus neoformans* and *Toxoplasma gondii*.

### Linearity

To evaluate the assay’s capability to accurately quantitate viral load for detected viruses, two linearity panels were generated from high-titer HCV positive serum and SARS-CoV-2 positive nasal swab samples. For both panels, the calculated linearity was 100% after running duplicates or triplicate replicates across a minimum of four 10-fold dilutions **(Supplementary Figure 2).** The absolute log_10_ deviation of calculated from expected viral loads was < 0.52 log_10_, which was favorable in comparison to the interquartile ranges for virus-specific qPCR assays between different laboratories^26^.

### Precision

We measured intra-assay precision by testing two PC and two NC samples within the same run using different barcodes across 20 runs and inter-assay precision by testing 20 PC and 20 NC samples using different barcodes across 20 separate runs. Essential agreement (EA) was 100% and intra- and inter-assay precision showed log-transformed coefficients of variation in reads per million of <10% and <30%, respectively **(Table 1)**.

### Inclusivity and Exclusivity

To evaluate the ability of the mNGS assay to detect a wide range of targets (inclusivity), we obtained commercially available culture supernatants from 17 respiratory viruses representing different sublineages and subspecies. Viruses were spiked into negative control matrix at concentrations ranging from 1.3 x 10^3^ to 1.2 x 10^7^ 50% tissue culture infective dose (TCID50) per mL **(Table 2)**. All 17 (100%) of 17 viruses in these contrived samples were correctly identified by mNGS assay at the sublineage or subspecies level. Additionally, we identified subtypes of rhinovirus and enterovirus from PCR-positive clinical samples that were not differentiated by multiplex RT-PCR (**Supplementary Figure 3A**). We also evaluated the ability of the mNGS assay to identify uncommon or rare viral pathogens associated with respiratory infections (n=8 virus-positive tracheal aspirate samples) or central nervous system (CNS) infections (n=4 cerebrospinal fluid samples) in severely ill hospitalized patients **(Table 2, Supplementary Figure 3B).** The assay detected 11 (100%) of 11 viruses in these samples. To assess the exclusivity of the mNGS assay, we spiked a previously established panel of 7 representative pathogenic organisms into NC matrix and analyzed multiple aliquots **(Table 1)**. Detected reads from non-viral pathogenic organisms did not result in any false-positive detections for viral pathogens. Although only 7 pathogens were tested, each of the key categories of organisms that are encountered in respiratory samples (DNA virus, RNA virus, Gram-positive bacterium, Gram-negative bacterium, fungus, mold, and parasite) were represented on the panel, and we have previously shown that results from this panel could be extrapolated for detection of other species within the same category^27^.

**Table 2.** Detection of a broad range of viruses in contrived samples.

| Contrived Sample type | Correctly identified Virus by mNGS assay |  |
| --- | --- | --- |
| Positive cerebrospinal fluid (CSF) spiked in negative matrix | Lymphocytic Choriomeningitis Virus (LCMV) |  |
|  | Herpes simplex virus 2 (HSV-2) |  |
|  | Varicella-zoster virus (VZV) |  |
|  | Herpes simplex virus 1 (HSV-1) and Epstein-Barr Virus (EBV) |  |
| Positive bronchoalveolar lavage (BAL) spiked in negative matrix | Parainfluenza Virus Type 4 | Parechovirus A |
|  | Influenza C Virus | Human Bocavirus |
|  | Primate Bocaparvovirus 1 | Coronavirus 229E |
|  | Coronavirus NL63 |  |
| Viral culture fluid spiked in negative matrix | Adenovirus | Coronavirus 229E |
|  | Coronavirus NL63 | Coxsackie Virus Type A1 |
|  | Echovirus | Human Metapneumovirus 16 |
|  | Influenza B Virus | Measles Virus |
|  | Mumps Virus | Parainfluenza Virus Type 2 |
|  | Parainfluenza Virus Type 3 | Parainfluenza Virus Type 4A |
|  | Parechovirus Type 1 | Rhinovirus A16 |
|  | Rhinovirus B14 | Rubella Virus |
|  | Influenza B Virus |  |

### Contamination, Interference, Matrix Effect and Stability

We evaluated potential cross-contamination between nearby sample wells and carryover contamination across successive runs from 10 SARS-CoV-2 high-titer clinical samples and 24 controls (cycle threshold, or C_t_ = 16-20) loaded in modified checkboard pattern (with at least one space between samples) on a 96-well plate, to mimic a single run on the Illumina NextSeq instrument. Only one cross-contamination event was observed, with a single SARS-CoV-2 read detected in one of th negative control wells at a subthreshold reporting leve. We also evaluated the effects of interference from human RNA, bacterial DNA, and potential interfering substances on mNGS assay performance. Hemolysis, lipids, bilirubin, and human genomic RNA spiked into PC matrix at concentrations of 0.1 – 100 µg/mL did not interfere with respiratory virus detection, but background DNA/RNA spiked into PC matrix at concentrations ≥1 x 10^7^ cells/mL resulted in failure to detect viruses due to high background. To evaluate the potential matrix effect from samples with high host background, we analyzed a total of 20 PCR-positive bronchoalveolar lavage (BAL) samples obtained from patients with symptomatic respiratory illnesses and patients who had routine bronchoscopy post-lung transplantation. Six (30%) of the BAL samples did not meet the sequencing quality control criteria (detection of MS2 phage and/or virus and at least 5 million preprocessed reads, **Supplementary Table 1**) and were excluded from analyses. Among the remaining valid samples (n=14), the assay successfully detected 92.9% (13/14) of the targeted pathogens. Of the 14 valid samples, six (43%) exhibited zero MS2 phage reads, suggesting an extremely high host background **(Supplementary Table 3)**. Finally, we evaluated mNGS assay stability; qualitative detection was not affected by keeping samples for up to 7 days at 4°C or subjecting the samples to 3 freeze/thaw cycles.

### Accuracy

To evaluate accuracy, 191 residual samples after routine clinical testing were obtained from the UCSF Clinical Microbiology Laboratory, including 110 virus-positive samples (103 NPS samples and 7 BAL fluids) from patients with acute respiratory infection, along with 81 virus-negative samples (52 NPS samples and 29 BAL fluids) **(Figure 4)**. As more than one target may be positive with mNGS and viral multiplex panel (VRP) testing using FDA-approved in vitro diagnostic (IVD) assays, sensitivity/specificity analyses were performed by assessing each result independently to assign true/false-positive/negative calls (see Methods for details). Compared to results from VRP RT-PCR testing, the mNGS assay exhibited 93.6% (103 of 110) sensitivity, 93.8% (76 of 81) specificity, and 93.7% (179 of 191) accuracy.

**Figure 4.**
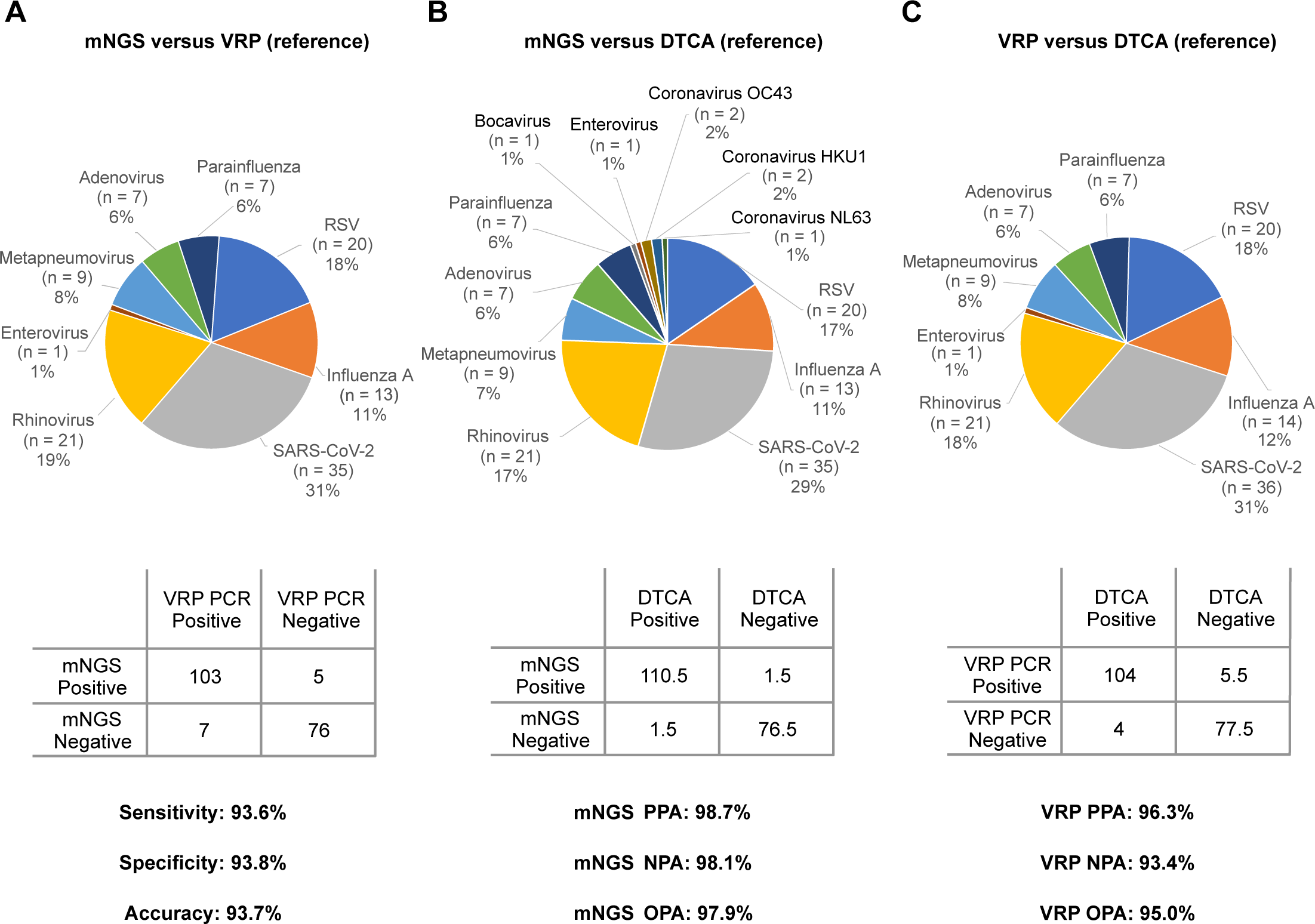
Accuracy evaluation for the mNGS assay. Pie charts and 2×2 contingency tables showing the distribution of detected viruses and performance metrics. **(A)** mNGS against VRP, **(B)** mNGS against DTCA, and **(C)** Clinical VRP against DTCA. VRP testing using FDA IVD assays includes detection of respiratory syncytial virus, parainfluenza viruses 1 to 3, metapneumovirus, rhinovirus/enterovirus, influenza A virus, and influenza B virus, and adenovirus. Discrepant samples that were mNGS-positive/VRP-negative or mNGS-negative/VRP-positive underwent orthogonal testing by targeted virus-specific PCR at the state public health laboratory and medical chart review for the most likely diagnosis by clinical adjudication. Abbreviations: mNGS, metagenomic next-generation Sequencing; PCR, polymerase chain reaction; VRP, viral respiratory panel; DTCA, discrepancy testing and clinical adjudication; PPA, positive percent agreement; NPA, negative percent agreement; OPA, overall percent agreement; RSV, respiratory syncytial virus; FDA, Food and Drug Administration; IVD, in vitro diagnostic.

Discrepancy testing and clinical adjudication (DTCA) of 14 mNGS positive-VRP negative samples using blinded chart review by two board-certified infectious diseases physician (PB and CYC) and orthogonal assays run by the California Department of Public Health Viral and Rickettsial Disease Laboratory confirmed the presence of 9 respiratory viruses missed by VRP, allowing them to be reclassified as true positives **(Table 3)**. Viruses detected by mNGS but not targeted by VRP were not considered false-positive results. In one case, while the original VRP and orthogonal PCR testing returned negative results, mNGS identified rhinovirus C with high confidence. A review of the viral sequences revealed 12 non-overlapping reads across the human rhinovirus C genome **(Supplementary Figure 4)**. Cross-contamination was ruled out, as no other sample in the sequencing batch tested positive for rhinovirus. A nucleotide BLAST (blastn) search confirmed sequences with high homology (95-98% identity) to known rhinovirus C strains **(Supplementary Data 1)**. Although the exact primer binding sites for the clinical RT-PCR assays used in the current study are unknown, we identified, for the rhinovirus C sample, the presence of mismatches in primer and probe regions from previously reported RT-PCR assays targeting the 5’-untranslated region (UTR)^28,29^ **(Supplementary Figure 4C),** which explained the detection by mNGS despite negative RT-PCR results.

**Table 3.** Discrepant mNGS Positive Results Compared to Original Viral Respiratory Panel Clinical Testing (n = 14 Samples)

| **Organisms Detected by Clinical VRP** | **Organisms Detected by mNGS** | **Results of Discrepancy Testing** | **Additional Data** | **Final Result of mNGS by DTCA** |
| --- | --- | --- | --- | --- |
| RSV | RSV<br>Rhinovirus B | (+) Rhinovirus |  | **TP** RSV<br>**TP** Rhinovirus |
| RSV | RSV<br>EVD68 | (+) Enterovirus |  | **TP** RSV<br>**TP** Enterovirus |
| RSV | RSV<br>Adenovirus C | (+) Adenovirus |  | **TP** RSV<br>**TP** Adenovirus |
| Adenovirus<br>Metapneumovirus | Adenovirus C<br>Metapneumovirus<br>Bocavirus | Bocavirus ND | Bocavirus mNGS reads are specific to the viral genus. Patient with clinical syndrome consistent with acute viral respiratory infection. | **TP** Adenovirus<br>**TP** Metapneumovirus<br>**TP** Bocavirus |
| ND | Coronavirus OC43 | Coronavirus OC43 ND | Detected by mNGS x 2 with high RPM and high coverage. No evidence of cross-contamination. Patient with clinical syndrome consistent with acute viral respiratory infection. | **TP** Coronavirus OC43 |
| ND | Coronavirus OC43 | (+) Coronavirus OC43 |  | **TP** Coronavirus OC43 |
| ND | Rhinovirus A | (+) Rhinovirus |  | **TP** Rhinovirus |
| ND | Rhinovirus A | (+) Rhinovirus |  | **TP** Rhinovirus |
| ND | Coronavirus HKU1 | Coronavirus HKU1 ND |  | **FP** Coronavirus HKU1 |
| ND | Coronavirus HKU1 | (+) Coronavirus HKU1 |  | **TP** Coronavirus HKU1 |
| ND | Parainfluenza 1<br>SARS-CoV-2 | (+) Parainfluenza 1<br>SARS-CoV-2 ND |  | **TP** Parainfluenza 1<br>**FP** SARS-CoV-2 |
| ND | Coronavirus NL63 | (+) Coronavirus NL63 |  | **TP** Coronavirus NL63 |
| ND | Rhinovirus C | Rhinovirus ND | Mismatches in the forward primer and probe sequences for rhinovirus/enterovirus PCR. No evidence of contamination. Patient with clinical syndrome consistent with acute viral respiratory infection. | **TP** Rhinovirus |
| ND | Coronavirus OC43 | NT |  | **NA** Coronavirus OC43 |

False positive results were only considered if the organism was included as a target for VRP. Each result was assessed independently, in case of multiple positive results (multiple organisms), the corresponding TP/FP call was divided by the number of organisms detected within each sample (e.g. 1 TP + 1 FP in a same sample = ½ TP + ½ FP). This allowed to keep the observation constant (see Methods for further details). (VRP) Viral respiratory panel; (DTCA) Discrepancy Testing and Clinical Adjudication (RSV) Respiratory Syncytial Virus; (EVD68) Enterovirus D68; (RPM) Reads per million; (ND) Not detected; (NT) Not tested; (TP) True positive; (FP) False positive; (NA) Not available because did not undergo DTCA.

Similarly, DTCA was performed on the 7 mNGS negative / VRP positive samples along with repeating the VRP assay (if possible, on a different instrument). This reassessment resulted in 5.5 samples being reclassified as true negatives (1 sample harbored two organisms adjudicated as one true negative and one false negative) **(Table 4)**. Compared to a composite standard that incorporates discrepancy testing and clinical adjudication, positive, negative, and overall predictive agreements of the mNGS assay were 98.7% (110.5 of 113), 98.1% (76.5 of 78), and 97.9% (187 of 191), respectively.

**Table 4.** Discrepant mNGS Negative Results Compared to Original Viral Respiratory Panel Clinical Testing (n = 9 Samples)

| **Organisms Detected By Clinical VRP** | **CT Value or MFI from Clinical VRP** | **Organisms Detected by mNGS** | **High Host Background?** | **Results of Discrepancy Testing** | **Discrepancy Testing Platform** | **Final Result of mNGS by DTCA** |
| --- | --- | --- | --- | --- | --- | --- |
| Influenza A<br>Parainfluenza 3 | NA | Influenza A | Y | (+) Influenza A<br>Parainfluenza 3 ND | MAGPX | **TP** Influenza A<br>**TN** Parainfluenza 3 |
| Influenza A and<br>SARS-CoV-2 | SARS-CoV-2 CT: 36.4 | Influenza A | Y | Influenza A NT<br>SARS-CoV-2 ND | Verigene | **TP** Influenza A<br>**TN** SARS-CoV2 |
| Influenza A<br>RSV | NA | ND | Y | Influenza A ND<br>(+) RSV B | GenMark | **TN** Influenza A<br>**FN** RSV |
| Influenza A and<br>SARS-CoV-2 | NA | SARS-CoV-2 | N | Influenza A ND<br>SARS-CoV-2 NT | MAGPX | **TP** SARS-CoV-2<br>**TN** Influenza A |
| Rhinovirus/Enterovirus | Rhinovirus/Enterovirus<br>MFI: 52 | ND | Y | Rhinovirus/Enterovirus ND | MAGPX | **TN** Rhinovirus/Enterovirus |
| Rhinovirus/Enterovirus | NA | ND | N | Rhinovirus/Enterovirus ND | MAGPX | **TN** Rhinovirus/Enterovirus |
| Rhinovirus/Enterovirus<br>Parainfluenza 2 | NA | Rhinovirus A | N | (+) Rhinovirus/Enterovirus<br>Parainfluenza 2 ND | MAGPX | **TP** Rhinovirus/Enterovirus<br>**TN** Parainfluenza 2 |
| Metapneumovirus | NA | ND | Y | (+) Metapneumovirus | GenMark | **FN** Metapneumovirus |
| RSV A | NA | ND | Y | RSV ND | MAGPX | **TN** RSV A |

(VRP) Viral respiratory panel; (MFI) Mean Fluorescence Units; (DTCA) Discrepancy Testing and Clinical Adjudication; (RSV) Respiratory Syncytial Virus; (ND) Not detected;
(Y) Yes; (N) No; (NA) Not available; (NT) Not tested; (TP) True positive; (FN) False negative

### Detection of divergent viruses

To benchmark the capability of the modified SURPI+ pipeline for detection of novel, highly divergent viruses *in silico*, we created a simulated sequencing output file containing many known human viruses of clinical and public health significance, including those with pandemic potential **(Figure 2B)**. We then took a higher-level set of taxonomic identifiers (species, genus, and/or family) corresponding to these viruses and removed all entries with these taxonomic identifiers from the SURPI+ 2019 reference dataset. Next, we used the SURPI+ pipeline to analyze the simulated sequencing file against both the original and “filtered” reference databases **(Figure 2C)**. In this analysis, 98.6% (69 of 70) of human viruses were detected at a sequencing depth of 100 reads per million (RPM) and 100% (70 of 70) at 1000 RPM based on homology to known animal or plant viruses. Of note, bunyaviruses pathogenic to humans, which are among the most divergent viruses, were still identified by translated nucleotide (amino acid) alignment to plant viruses (for example, detection of Venezuelan equine encephalitis virus based on homology to vanilla latent virus in **Figure 2C, “VEEV”**).

## Discussion

We validated a clinical mNGS assay in a CLIA laboratory as a Laboratory Developed Test (LDT) for agnostic viral respiratory pathogen detection intended to aid in patient diagnosis and public health surveillance. Our main goal was to develop, optimize, and streamline a protocol for respiratory viral mNGS testing that could be deployed and run routinely in clinical or public health laboratories. The mNGS assay developed here has favorable performance characteristics compared to clinical VRP testing, including a limit of detection of ∼500 copies/mL, viral load quantification with 100% linearity, and sensitivity, specificity, and accuracy ranging from 93.6 – 93.8%. However, in contrast to targeted assays such as VRP, the mNGS assay is capable of detecting, in principle, all known as well as novel viral pathogens in respiratory samples. In addition, mNGS assay performance was found to be superior to VRP (97.9% versus 95.0% overall agreement) after discrepancy testing and clinical adjudication. Following completion of the validation, our assay received breakthrough device designation from the US Food and Drug Administration (FDA) in August of 2023. Widespread implementation of highly accurate, rapid mNGS assays such as this, with enhanced capacity to detect novel viruses, will support robust preparation for and rapid response to the next viral pandemic.

Speed is a critical factor for diagnosis of respiratory infections, especially in critically ill patients with lower respiratory involvement and in outbreak investigations of novel or emerging viruses with pandemic potential. We also aimed to develop an assay that could be deployable widely in clinical and public health laboratories. Thus, we optimized many of the steps of the mNGS assay and moved the key RNA/cDNA library preparation step to an automated platform, the MagicPrep NGS system (Tecan Genomics, Inc., Männedorf, Switzerland). We further demonstrated that sequencing can be performed on the Illumina MiniSeq using the Rapid Reagent Kit for a faster 5-hour turnaround time or on the Illumina NextSeq 550Dx using the Mid-Output Reagent Kit for a 13-hour turnaround time, depending on laboratory needs and priorities. All together, these modifications resulted in an assay with a turnaround time of 14-24 hours and ∼2 hours of hands-on technician time.

Orthogonal testing and clinical adjudication performed on discordant results demonstrated that the VRP assay is an imperfect gold standard on which to judge mNGS performance. The mNGS assay was able to not only detect uncommon infections from viruses not covered on existing VRP panels, but also, in multiple cases, detect viruses that would in principle be detectable by VRP but tested negative. Unlike VRP, mNGS does not rely on specific primers or probes and is thus less susceptible to primer failure due to viral evolution, as demonstrated with the mNGS positive and VRP negative rhinovirus case presented here, and which can result in decreased assay sensitivity or false negative results due to viral mutation, which is an inevitable feature of SARS-CoV-2 and many other RNA viruses^30^. Notably, a previous study evaluating the usefulness of published PCR primers in detecting rhinovirus infection reported that none of the published rhinovirus-specific PCR primer pairs could detect all human rhinoviruses in 101 genotyped clinical specimens^31^. In addition, the broader sampling of the viral genome by mNGS may result in increased sensitivity of virus detection compared to VRP due to increased robustness to variability in the relative levels of viral gene expression by infected cells^32^. Most of the false-negative mNGS samples were confirmed as true negative after chart review and repeating the VRP assay. Most likely, these represented false-positive results during the original VRP run, given the high cycle thresholds (>36), suggesting low viral titers, or samples that had degraded over time and/or after multiple freezing and thawing cycles.

In the study, we used several approaches to demonstrate the capacity of the mNGS assay to identify novel and/or emerging viruses with divergent genomes. The assay was successful in detecting uncommon and unusual viral pathogens associated with both severe respiratory infections (bronchoalveolar lavage fluid) and central nervous infections (CSF spiked into respiratory sample matrix). mNGS testing also enabled subtyping of specific viral strains with increased virulence, such as enterovirus D68, which has been linked to acute flaccid myelitis in children^33,34^, and rhinovirus C, which has been associated with invasive pulmonary and bloodstream infection in immunocompromised patients^35,36^. Importantly, the mNGS assay was also able to detect DNA viruses, such as adenovirus and bocavirus, in both clinical and contrived samples, despite the incorporation of DNase treatment in the protocol. Detection of DNA viruses is presumably based on detection of transcribed viral mRNA in infected cells, although may also enabled by incomplete DNA digestion from.the DNase enzyme.

To evaluate the capacity for mNGS testing using a modified SURPI+ computational pipeline to identify novel viruses, we performed an *in silico* analysis of a contrived metagenomic dataset consisting of reads from the genomes of human viruses of pandemic potential spiked into background using a reference database depleted of all known human viral sequences. This analysis was done to simulate whether “novel” human viruses with pandemic potential could be identified based on homology to known plant and animal viruses. All 70 of the human viral pathogens tested were successfully identified, including those with only remote homology to other viruses. Indeed, chikungunya virus, in the *Alphavirus* genus of the *Togaviridae* family, was only identified (after removal of all alphaviruses) because of distant homology to vanilla latent virus in the family *Alphaflexivirdae*. Notably, alphaflexiviruses contain a distinct lineage of alphavirus-like replication proteins that lack a recognized protease domain^37^.

Our validation study has limitations. First, only a limited number and diversity of clinical samples harboring rare or unusual respiratory viruses were available, necessitating the addition of contrived samples to our validation study. Second, we did not formally prove that the mNGS assay would be able to detect a novel, sequence-divergent virus, but instead demonstrated the ability of the test to detect such a virus using an *in silico* analysis, an approach which has been used in previous studies to benchmark mNGS bioinformatic pipelines for viral pathogen discovery^38,39^. Finally, we did not address the utility of the mNGS assay for routine diagnosis in patients with unexplained infections, or for outbreak surveillance in public health, which will likely require future prospective clinical and/or epidemiologic investigation.

## Resource availability

### Lead Contact

Further information and requests for resources and reagents should be directed to and will be fulfilled by the Lead Contact, Charles Chiu.

### Materials Availability

This study did not generate any new reagents.

## Data and Code Availability

Human-subtracted raw sequence data were submitted to the Sequence Read Archive (SRA) database. (BioProject accession number <u>PRJNA1084017</u> and umbrella BioProject accession number PRJNA171119). Sequence metadata, custom scripts and code for data analyses and visualization are available in a Zenodo data repository (https://doi.org/10.5281/zenodo.10553379).

## Methods details

### Human Sample Collection

Laboratory-confirmed virus-positive NP or BAL samples were retrieved from the UCSF Clinical Microbiology Laboratory and stored in a biorepository according to protocols approved by the UCSF Institutional Review Board (protocol no. 11-05519) until processed. Residual samples were required to meet minimal sample handling, storage, and volume requirements for inclusion in our study. Included samples were stored at 4°C for <24 hr prior to being de-identified, aliquoted, and stored in -80°C freezer prior to mNGS processing, thus undergoing one freeze-thaw cycle.

### Inclusion and Ethics

All residual samples meeting minimal requirements were included in the study. Samples were de-identified prior to processing.

### External controls preparation

External positive control (PC) was prepared by spiking a pooled negative nasal swab matrix with a commercially available reference material, the Accuplex Verification Panel (SeraCare, Milford, MA). This panel consists of a mixture of non-infectious SARS-CoV-2, influenza A, influenza B, and RSV genomes encapsidated in a synthetic protein coat to mimic the structure of a viral capsid. This PC material was “spiked in” at a titer of approximately 10^4^ copies/mL for each virus control, which is 1–2 logs higher than the estimated limit of detection of the assay (∼500 copies/mL). The negative matrix was prepared by pooling nasopharyngeal swab samples from asymptomatic individuals and was used as an external negative control (NC).

### Nucleic acid extraction

500 µL of NP swab or BAL fluid was centrifuged at 16,000 x g for 10 minutes. The MagMAX™ Viral/Pathogen II (MVP II) Nucleic Acid Isolation Kit (Thermo Fisher Scientific, Waltham, MA) and the KingFisher™ Flex Purification System with a 96 deep-well head (Thermo Fisher Scientific, Waltham, MA) were used for total nucleic acid extraction. This protocol was modified to include DNase treatment as a host depletion step during extraction. Bacteriophage MS2 (Zeptometrix, Buffalo, NY) was added to all samples including the negative control as an internal qualitative control.

### Library preparation and sequencing

Simultaneous reverse transcription of purified RNA, spiked in with ERCC RNA controls (Invitrogen, Waltham, MA), and ribosomal RNA (rRNA) depletion were carried out using NEBNext® Ultra™ II RNA First Strand Synthesis Module (New England Biolabs, Ipswich, MA) and QIAseq FastSelect-rRNA HMR Kit (Qiagen, Germantown, MD), respectively, followed by second strand cDNA synthesis using Sequenase™ Version 2.0 DNA Polymerase (Thermo Fisher Scientific, Waltham, MA). Complementary DNA (cDNA) was purified using AMPure XP beads (Beckman Coulter, Brea, CA) and loaded on the MagicPrep NGS instrument (Tecan Genomics, Inc., Männedorf, Switzerland) to undergo end-repair, adapter ligation and barcoding, amplification (25 cycles) and purification. Libraries were quantified and normalized using the Qubit dsDNA HS Assay (Thermo Fisher Scientific, Waltham, MA) on the Qubit Flex (Thermo Fisher Scientific, Waltham, MA). Final pooled libraries were sequenced as single-end reads on either the Illumina (San Diego, CA) MiniSeq using the Rapid Reagent Kit (100 cycles) or on the Illumina NextSeq 550 using the Mid-Output or High-Output Kit (150 cycles).

### Bioinformatics

The SURPI+ computational pipeline, run as a container (v1.0.0) on either a secure server or cloud infrastructure, was used for identification of respiratory viral pathogens from mNGS data. Reads were preprocessed by trimming of adapters and removal of low-complexity and low-quality sequences, followed by computational subtraction of human reads. The Scalable Nucleotide Alignment Program (SNAP)^40^ nucleotide aligner was run using an edit distance of 16 against the National Center for Biotechnology Information (NCBI) nucleotide (NT) database (March 2019, with inclusion of the SARS-CoV-2 WuHan-Hu-1 genome accession number NC_045512) filtered to retain only viral reads. The pipeline was modified to include “tagging”, or annotation, of entries from reference sequences that constitute a subset of the NCBI NT database, such as FDA-ARGOS^22^. Note that the FDA-ARGOS database, while quality controlled and regulated, contains only 1,428 microbial strains, the majority of which are bacterial. It had also not been updated with recent viruses such as SARS-CoV-2; thus, we did not detect any reads matching to viral genomes in this study. The pipeline was also modified to include optional de novo assembly and protein sequence alignment using SPAdes ^41^ and DIAMOND ^42^, respectively. Coverage maps were automatically generated by mapping reads classified by SURPI as viral to the most likely reference genome.

Quality control metrics for the assay were based on those previously established for cerebrospinal fluid^20^, and include a minimum of 5 million preprocessed reads per sample, >75% of data with quality score >30 (Q>30), and successful detection of the 4 respiratory viruses in the PC and the internal spiked MS2 phage control. A threshold criterion of ≥3 non-overlapping viral reads aligning to the target viral genome was considered a positive detection.

### Evaluation of mNGS analytical performance characteristics

The automated standard operating procedures and sequencing runs for these clinical validation studies were performed by a state-licensed clinical laboratory scientist. LoD was determined for each of the four representative organisms in the PC by probit analysis using a series of dilutions ranging from 100 to 5,000 copies/mL, with 10 to 40 replicates at each concentration. Linearity was demonstrated by plotting the standard curve. To validate the quantification accuracy using the ERCC and the positive control, we serially diluted an HCV positive plasma to known concentration ranging from 4 x 10^6^ to 4 x 10^3^ copies/mL in triplicates. We then compared the quantitative measure to the known measure. Precision was determined using repeat analysis of two PC and two NC samples across 20 runs (intra-assay reproducibility) and by testing 20 PC and 20 NC across 20 separate runs (inter-assay reproducibility). To assess inclusivity, commercially available cultured supernatants were obtained to assess the assay’s ability to detect the intended targets. Each of the 17 respiratory viruses, titers ranging from 1.3 x 10^4^ to 1.2 x 10^8^ TCID50/mL, were spiked into the negative control matrix at 1:10 dilutions. These viruses represented known sublineages and subspecies and we evaluated their identification by our assay. We also tested samples of confirmed virus-positive BAL (n=7) and CSF samples (n=4) spiked into negative matrix to evaluate the detection of unusual viruses. To assess the exclusivity of the mNGS assay, we spiked a previously established mixture of seven representative pathogenic organisms to verify the false positive detection for viral pathogens. We evaluated cross-contamination between adjacent sample wells and carryover contamination across successive runs from samples with high viral loads. Interference was determined using PC spiked with known amount of hemolytic blood, lipids, bilirubin, human RNA, bacterial DNA/RNA. The effect of mucus in BAL positive fluid was also assessed. Stability was determined by keeping samples for up to 7 days at 4°C or subjecting the samples to 3 freeze/thaw cycles. Accuracy was determined using 191 clinical samples comprising 110 virus-positive samples (103 NPS samples and 7 BAL fluids) from patients with acute respiratory infection, along with 81 virus-negative samples (52 NPS samples and 29 BAL fluids). Samples were obtained from patients at the University of California, San Francisco (UCSF). The viral RT-PCR comparator assays that were used include the Genmark ePlex (Carlsbad, CA), Luminex NxTAG (Austin, TX), and/or Luminex Verigene RP Flex Respiratory Pathogen Panels. mNGS results were compared with original clinical testing and then with a composite reference standard including discrepancy testing and clinical adjudication. In the second comparison, when results were discordant, orthogonal testing was performed using a different instrument or an independent CLIA laboratory (the California Department of Public Health) in addition to clinical adjudication to reclassify mNGS results. The second comparison was reported as PPA and NPA, as selective discrepancy testing can bias sensitivity and specificity results.

### Orthogonal discrepancy testing at the California Department of Public Health

Specimens were tested by real-time PCR based on CDC protocols using a viral respiratory panel, an unpublished CDPH laboratory-developed test (LDT). Viruses that can be detected by this panel include human metapneumovirus, respiratory syncytial virus, adenovirus, parainfluenza virus (types 1, 2, 3, and 4), enterovirus/rhinovirus, and human coronaviruses 229E, OC43, NL63, and HKU1.

### *In* silico analysis for identification of novel and/or divergent viruses using the SURPI+ pipeline

To measure accurate detection of novel and/or divergent viruses, an *in silico* analysis was performed. Representative viral reference genomes corresponding to outbreak viruses of clinical and public health significance with pandemic potential were retrieved from the NCBI GenBank database, partitioned into non-overlapping segments, and then randomly sampled and spiked *in silico* into a negative nasal swab matrix sequencing library. We then took a higher-level set of taxonomic identifiers (species, genus, and/or family) corresponding to these viruses and removed all entries with these taxonomic identifiers from the SURPI+ reference dataset. Next, we used the SURPI+ pipeline to analyze the simulated sequencing file against both the original and “restricted reference” databases and evaluated the performance of the pipeline in detecting “simulated” novel and/or divergent viruses that lacked a reference sequence.

### Statistical analyses

Sensitivity and specificity analyses were performed as follows: as more than one target may be positive with mNGS and VRP, each result was independently assessed in every sample and true/false-negative/positive were accordingly assigned to each result. However, the total number of observations was kept constant (one sample = one observation = 1). For instance, in the case a test detected two organisms, namely the real culprit pathogen and a contaminant, the former was assigned 0.5 true-positive (TP) and the latter 0.5 false-positive (FP), in order as their sum was always equal to 1. In addition, as we used VRP as a comparator which includes a limited number of targets, mNGS positive-VRP negative results that were not a target for the VRP were not considered as false-positive results.

Statistical analyses were performed using scipy (version 1.5.3) and rstatix (version 0.7.0) packages as implemented in Python (version 3.7.12) and R (version 4.0.3), respectively. Probit regression analyses were done using scipy (version 1.5.3), numpy (version 1.19.1) and statsmodels (version 0.12.2) as implemented in Python software (version 3.7.12).

## Data Availability

Data and Code Availability
Human-subtracted raw sequence data were submitted to the Sequence Read Archive (SRA) database. (BioProject accession number PRJNAXXX and umbrella BioProject accession number PRJNA171119). Sequence metadata, custom scripts and code for data analyses and visualization are available in a Zenodo data repository (https://doi.org/10.5281/zenodo.10553379).
 

https://doi.org/10.5281/zenodo.10553379

## Acknowledgments

We thank the staff at the UCSF Clinical Microbiology Laboratory for help in collecting nasopharyngeal swab and bronchoalveolar lavage fluid samples. This work was financially supported in part by BARDA EZ-BAA award 75A50122C00022 (C.Y.C.), US CDC grants 75D30122C15360 and 75D30121C12641 (C.Y.C.), Abbott Laboratories (C.Y.C.), and the Chan-Zuckerberg Biohub (C.Y.C.). The funders had no role in the design and conduct of the study; collection, management, analysis, and interpretation of the data; preparation, review or approval of the manuscript; and decision to submit the manuscript for publication.

## Disclaimer

The content of this paper is solely the responsibility of the authors and does not represent the official views or opinions of the National Institutes of Health, Kaiser Permanente, California Department of Public Health or the California Health and Human Services Agency. Use of trade names and commercial sources is for identification only and does not imply endorsement by the California Department of Public Health or the California Health and Human Services Agency.

## Author contributions

J. Tan, V.S., D.S., and C.Y.C conceived and designed the study. J. Tan, V.S., D.S., N.S., A.F., H.J.H., J.N., M.O., N.B., J. Tang, D.I., B.F., H.R., M.H., C.M., D.A.W., and C.Y.C coordinated the sequencing efforts and laboratory studies. J. Tan, A.C., H.C., and S.Y. processed samples. J. Tan, V.S., D.S., E.K., A.C., H.C., S.Y., M.D.L., P.B., and C.Y.C. analyzed data. J. Tan, N.S., A.F., J.N., M.O., P.M.M., and C.L. collected samples. J. Tan, V.S., E.K., P.B., M.D.L and C.Y.C. wrote the manuscript. J. Tan, V.S., E.K., P.B., and C.Y.C. prepared the figures. J. Tan, V.S., D.S., E.K., N.S., A.F., H.J.H., J.N., M.O., N.B., J. Tang, D.I., B.F., H.R., M.H., D.A.W., P.M.M., C.R.L., M.D.L., P.B., and C.Y.C edited the manuscript. J. Tan, V.S., E.K., M.D.L., P.B., and C.Y.C. revised the manuscript. All authors read the manuscript and agree to its contents.

