## Supplementary Material for "Laboratory validation of a clinical metagenomic next-generation sequencing assay for respiratory virus detection and discovery"

**Supplementary Table 1. Evaluation and interpretation of human background based on detection of MS2 phage and/or viral reads.**

| **Result*** | **Interpretation** |
| --- | --- |
| MS2 phage DETECTED; viral pathogen DETECTED | Results are valid. |
| MS2 phage NOT DETECTED; viral pathogen DETECTED | Results are valid. |
| MS2 phage DETECTED; viral pathogen NOT DETECTED | Results are valid. |
| MS2 phage NOT DETECTED; viral pathogen NOT DETECTED | Results are invalid. |

* All possible outcomes regarding the validity of mNGS testing based on detection of MS2 and/or a viral pathogen in the clinical sample.

**Supplementary Table 2. Evaluation of human background based on ERCC RNA spike-in quantifications results.**

| **IC RPM Ratio Range*** | **Interpretive Comment** |
| --- | --- |
| >5 | Due to very low host background, estimated limits of detection in this clinical sample are approximately 1 log lower than the stated limits of detection for this test. |
| 0.5 – 5 | None |
| 0.05 – 0.5 | Due to moderate host background, estimated limits of detection in this clinical sample are approximately 1 log higher than the stated limits of detection for this test. |
| 0.005 – 0.05 | Due to high host background, estimated limits of detection in this clinical sample are approximately 2 logs higher than the stated limits of detection for this test. |
| 0 – 0.005 | Due to very high host background, estimated limits of detection in this clinical sample are approximately 3 logs higher than the stated limits of detection for this test. |

* $IC RPM ratio =\frac{ERCC RPM sample}{ERCC RPM NC}, for RPM NC >0, or 1 for RPM NC = 0$. Abbreviations: ERCC, External RNA Controls Consortium; IC, internal control; NC, negative control; RPM, reads per million.

**Supplementary Table 3. Interference study results for mucoid BAL.**

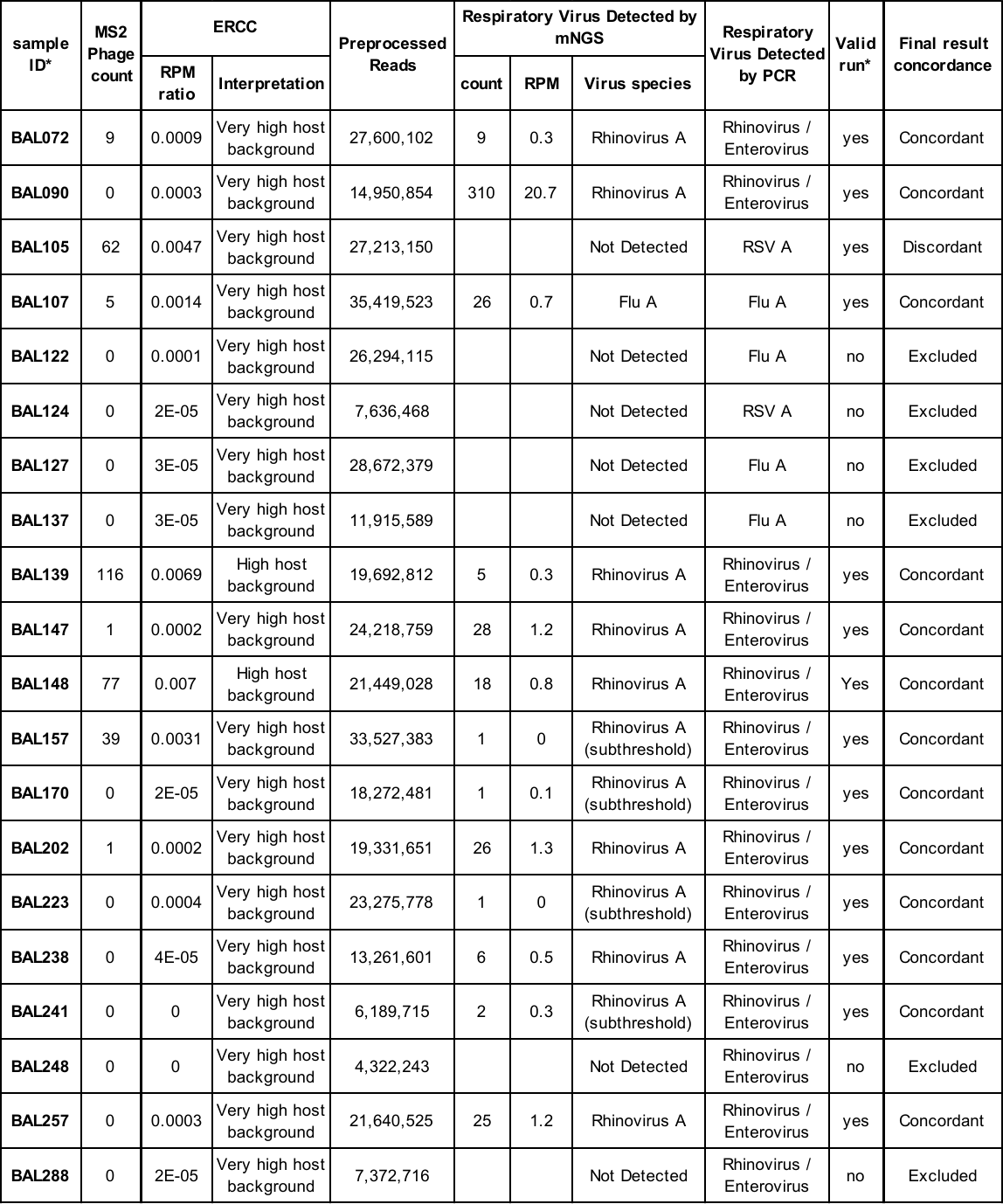

*Analysis of 20 PCR-positive bronchoalveolar lavage samples to evaluate the potential matrix effect from samples with high host background; invalid runs were excluded from the analysis. Abbreviations: ERCC, External RNA Controls Consortium; ST, subthreshold; BAL, bronchoalveolar lavage; RPM, reads per million.

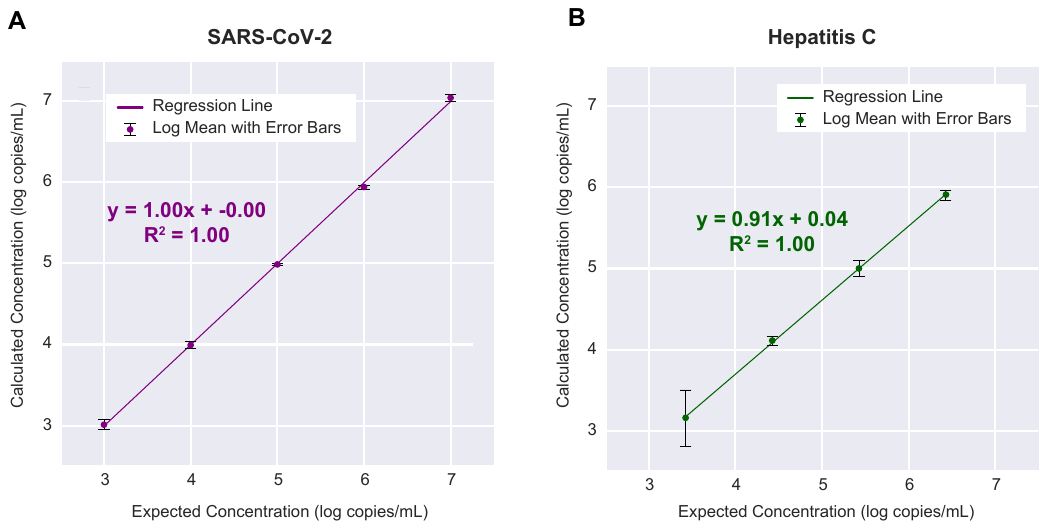

**Supplementary Figure 1.** **Evaluation of linearity and viral load quantification for the mNGS assay. (A)** A quantified SARS-CoV-2 PCR-positive nasopharyngeal swab from a patient with COVID-19 was serially diluted in donor nasal swab matrix and tested across 4 log_10_ dilutions. **(B)** A quantified HCV PCR-positive plasma sample from a patient with hepatitis C infection was serially diluted in donor plasma and tested across 4 log_10_ dilutions. At each dilution, the calculated mean concentration from three replicates is plotted against the expected concentration on a log scale, and the R^2^ correlation coefficient is determined by linear regression.

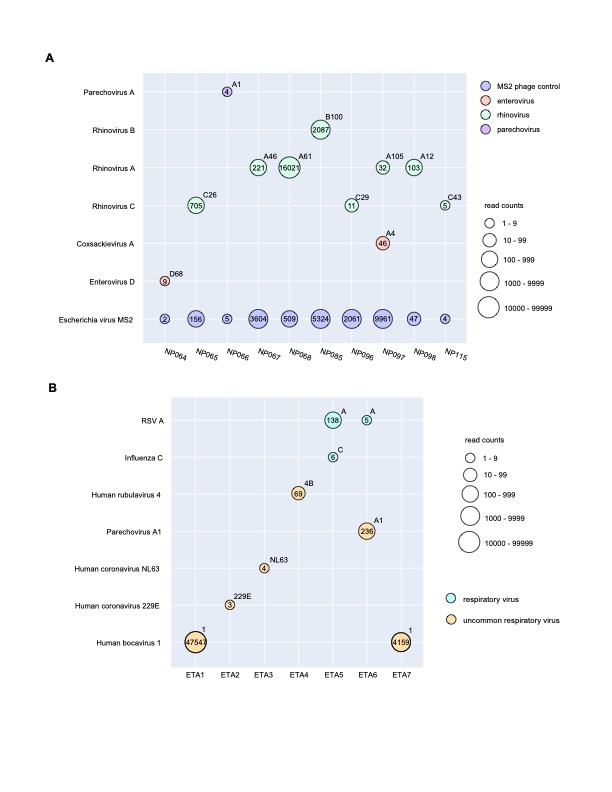

**Supplementary Figure 2. Demonstration of inclusivity and clinical use cases for the mNGS assay. (A)** Genotyping of rhinovirus and enterovirus subtypes from PCR-positive nasal swab samples. Conventional clinical multiplex RT-PCR tests do not distinguish between rhinoviruses and enteroviruses, nor are they able to subtype more pathogenic strains such as rhinovirus C or enterovirus D68 in association with acute flaccid myelitis^1,2^. **(B)** Detection of uncommon or rare viral pathogens causing respiratory infections in critically ill mechanically ventilated hospitalized patients. The circles correspond to detected viruses, and are color-coded by virus and scaled by read counts. For each detected virus, the read count is shown in the circle, while the identified genotype after SURPI+ pipeline is shown in the upper right quadrant. Abbreviations: ETA, endotracheal aspirate.

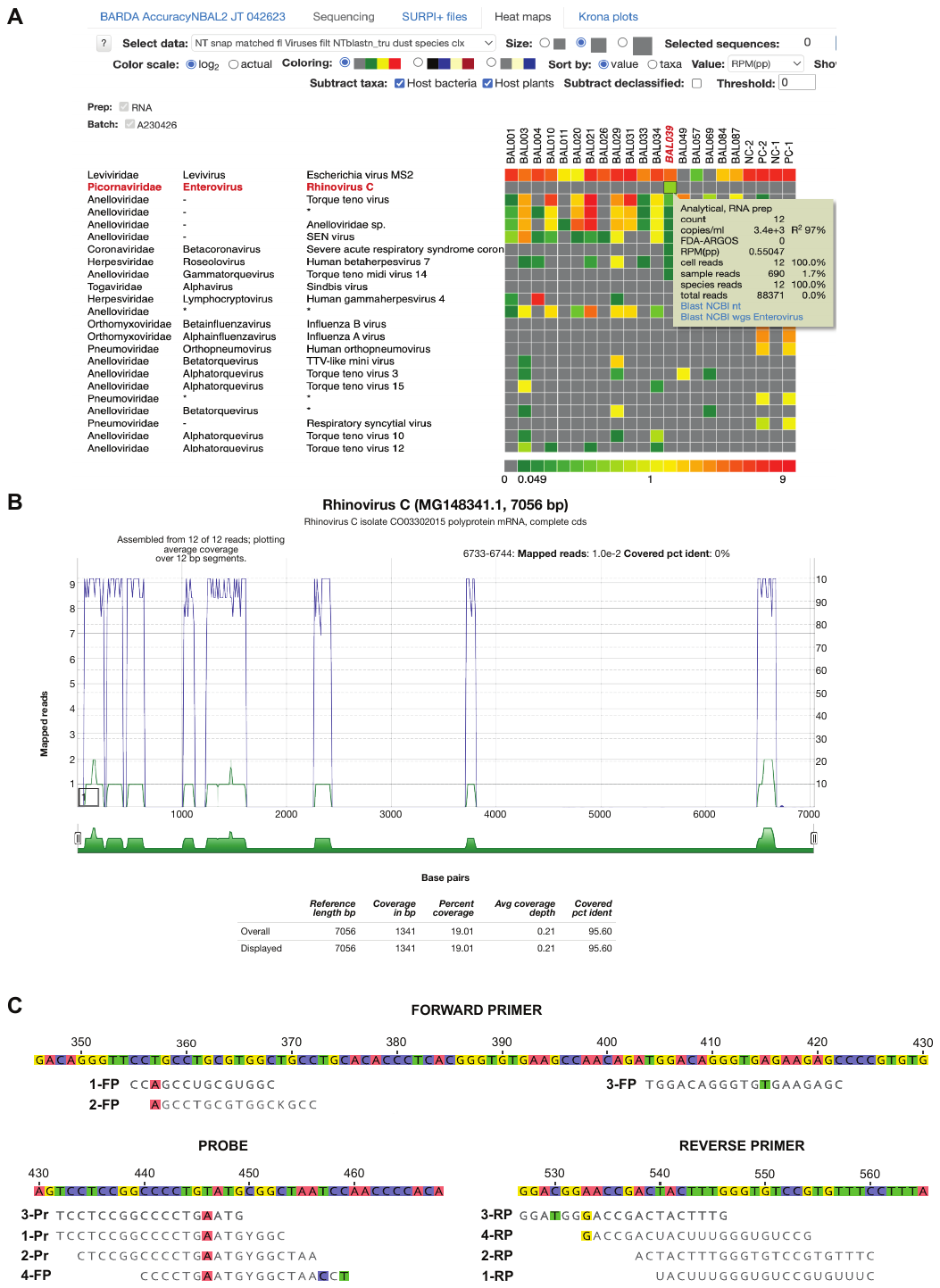
**Supplementary Figure 3.** **In-depth analysis of a rhinovirus C detection by mNGS that was discrepant with RT-PCR. (A)** A heat map generated from SURPI+ analysis shows 12 reads aligning to rhinovirus C from a single sample, excluding the possibility of cross-contamination. Each column denotes a clinical sample, while each row corresponds to a taxonomic identification at the species, genus, or family level. The asterisks refer to “declassification” of reads from one level to the next higher taxonomic level (for example, from species to genus). **(B)** A coverage map shows that the 12 reads span the genome of the most closely matched rhinovirus C genome in the reference database identified by SURPI+ (accession number MG148341.1) without overlap, with coverage of 19% of the ~7,000 base pair (bp) genome. **(C)** Several mismatches in the primer and probe sequences from published RT-PCR assays targeting the 5’-untranslated region (5’-UTR) are observed when compared to the viral mNGS reads, providing a likely explanation for the discrepant mNGS and RT-PCR results. The four assays are labeled 1 through 4 and correspond to Lu, et al.^3^ (1), Tapparel, et al.^4^ (2), Gunson, et al.^5^ (3), and Steininger, et al.^6^ (4). The mismatched nucleotides are highlighted with a background color. Note that the assay from Steininger, et al. does not include a probe. Abbreviations: FP, forward primer; Pr, probe. RP, reverse primer.
